# Global association of obesity and COVID-19 death rates

**DOI:** 10.1101/2021.03.09.21253222

**Authors:** Mary L. Adams

## Abstract

**Importance:** COVID-19 was responsible for an enormous global death toll with large variation among countries.

**Objective:** To examine the possible impact of obesity on COVID-19 death rates.

**Design:** Measure associations between obesity rates in 2016 and COVID-19 deaths/million population through 2/25/2021, across countries.

**Setting:** Global

**Participants:** 167 countries for which obesity and death data were available, grouped by population size, with multiples of 10 countries in each of 8 groups plus a group including all 57 countries with obesity rates <15%.

**Outcome and measures:** Using Excel, COVID-19 deaths/million were regressed on the obesity rate for each country, based on obesity being a key factor in COVID hospitalizations and deaths. Using the least squares formula for the best fit for each model, R^2^, components of the formula, and the percentage of world population represented, were recorded for each group.

**Results:** Obesity rates ranged from 2.1% to 37.9% and death rates ranged from 0.4/million to 1,892/million for groups representing up to 91% of global population. Results for the 8 population groups had R^2^ from 0.30 to 0.90 with slopes of the fitted line ranging from 27.9-51.0. Countries with obesity rates <15% had consistently low death rates (≤233/million), R^2^ of 0.003 and slope of the line=1.01.

**Conclusions:** For most countries about one-third of the difference in COVID death rates was due to obesity while in countries with obesity <15%, consistently low death rates were not associated with obesity. Reduced obesity rates could potentially have lowered the COVID death toll.

## 1. Background

Globally, the COVID-19 death toll varies from 0.4/million population in Viet Nam to 1,892/million in Belgium and the reason for such wide variation is unclear. Early in the pandemic, it was noted that people who reported certain underlying health conditions were more likely to die from COVID-19 (1,2). Initial US estimates of those likely to develop complications or die from COVID were based on data from China where the pandemic started and where obesity rates are low. Underlying conditions used for those estimates were hypertension, cancer, asthma, chronic obstructive pulmonary disease, diabetes, and cardiovascular disease (1,2). Once it was noted that hospitalized US adults were also more likely to be obese (3), estimates of US adults with underlying conditions were updated (4). Cancer was removed as an underlying condition and obesity was substituted, with the result that US estimates of adults with any of the underlying conditions increased from 45.4% to 56.0 % (4). Including obesity along with chronic diseases as a condition related to COVID deaths appears to blur the usual distinction between chronic and infectious diseases and suggested potential studies. The objective for this study was to analyze the association of obesity with death rates to estimate the contribution of obesity to the variation in COVID death rates among countries.

## 2. Methods

### 2.1. Study design & setting

The design involved regressing COVID death rates (as of February 25, 2021) (5) on obesity rates (6), by country. Participants were 167 countries with data for both obesity and deaths. Countries were grouped by population size into multiples of 10 countries in each of 8 groups, which included 90.9% of the world population. Results were compared with 57 countries with obesity rates <15%.

### 2.2 Analysis

Using Excel, equations for linear least squares trend lines and estimates of R^2^ were made from regression of COVID deaths/million population on obesity rates for each group. Using the least squares formula for the best fit for each model R^2^, components of the formula, and the percentage of world population represented (for population groups), were noted for each group.

## 3. Results

Obesity rates across the 167 countries ranged from 2.1% in Viet Nam to 37.9% in Kuwait. Death rates ranged from 0.4/million in Viet Nam to 1,892/million in Belgium. Figure 1. shows the graph with trendline and data points, R^2^ of 0.356, and formula for the line for the 60 most populous countries representing 87.6% of the world population. This is typical for the 8 graphs based on population. Figure 2 shows results for the 57 countries with obesity rates <15% with R^2^=0.0033 and a slope=1.01; all death rates are ≤233/million. A summary of the results for the 8 population groups plus results for all 167 countries is shown in Table 1. R^2^ for the 8 groups representing up to 90.9% of the population, ranged from 0.30 to 0.90 and slopes of the fitted least squares line ranged from 27.9-51.0. Doubling the number of countries to include all 167, but adding only 10% of world population, lowered R^2^ to 0.20 and the slope to 26.4.

**Table 1.** Summary of results for association between obesity and COVID death rates/million, adding countries to each group in order of highest population.

| Number in<br>group | $R^2$ | Fitted equation | | % Total pop |
| --- | --- | --- | --- | --- |
|  |  | Slope | +/- |  |
| 10 | 0.905 | 51.0 | -263.2 | 58.0 |
| 20 | 0.539 | 34.1 | -111.6 | 70.5 |
| 30 | 0.494 | 37.2 | -107 | 76.6 |
| 40 | 0.351 | 30.0 | -47.5 | 81.3 |
| 50 | 0.322 | 28.5 | -92 | 85.0 |
| 60 | 0.356 | 30.0 | -110 | 87.6 |
| 70 | 0.356 | 29.5 | -116 | 89.5 |
| 80 | 0.304 | 27.9 | -94.8 | 90.9 |
| 167 | 0.203 | 26.4 | -62.6 | ~100 |

**Figure 1 legend:**
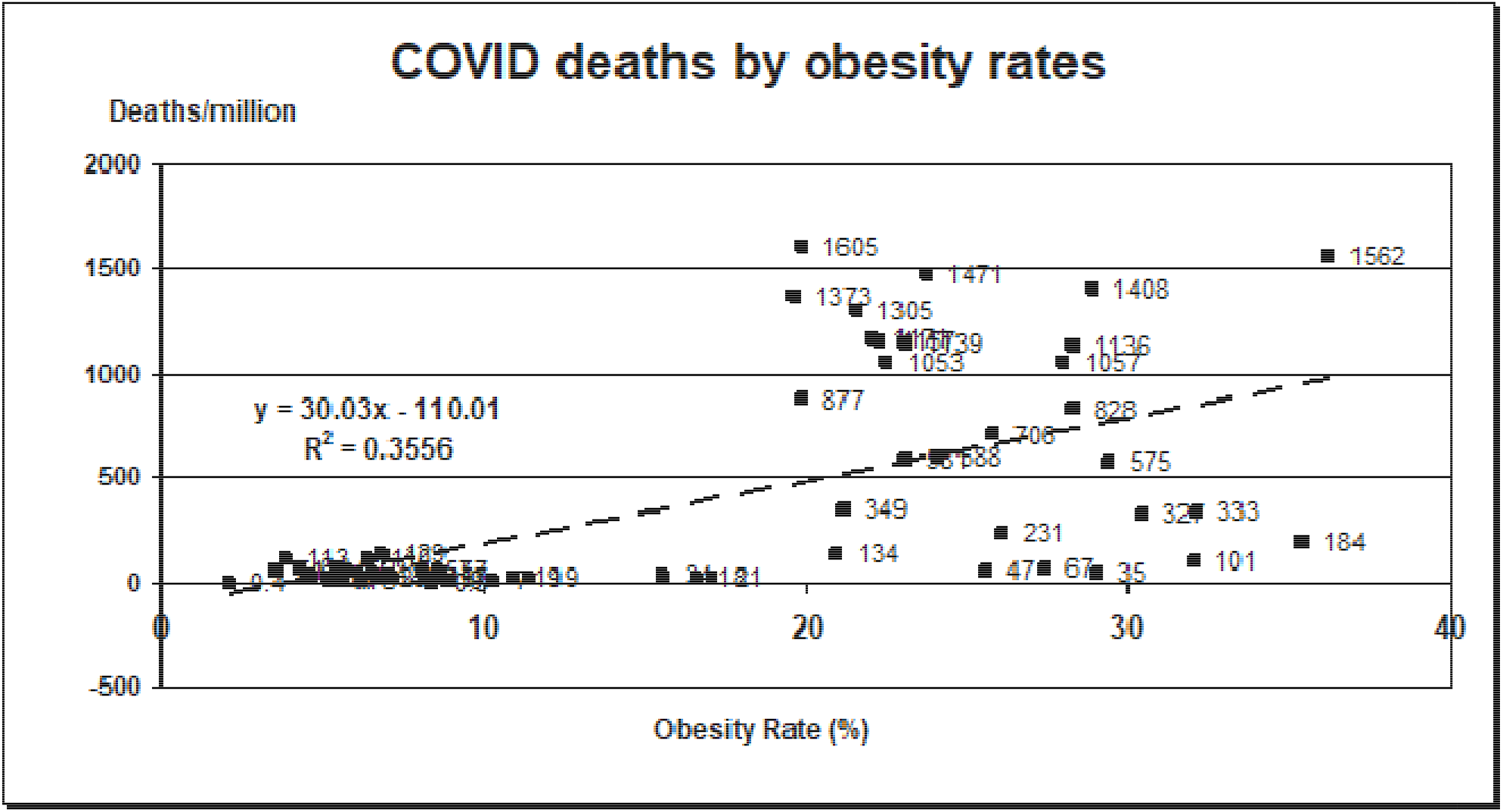
Association of 2016 obesity rates and COVID-19 deaths/million population for the 60 most populous countries representing 87.6% of the world population.

**Figure 2 legend:**
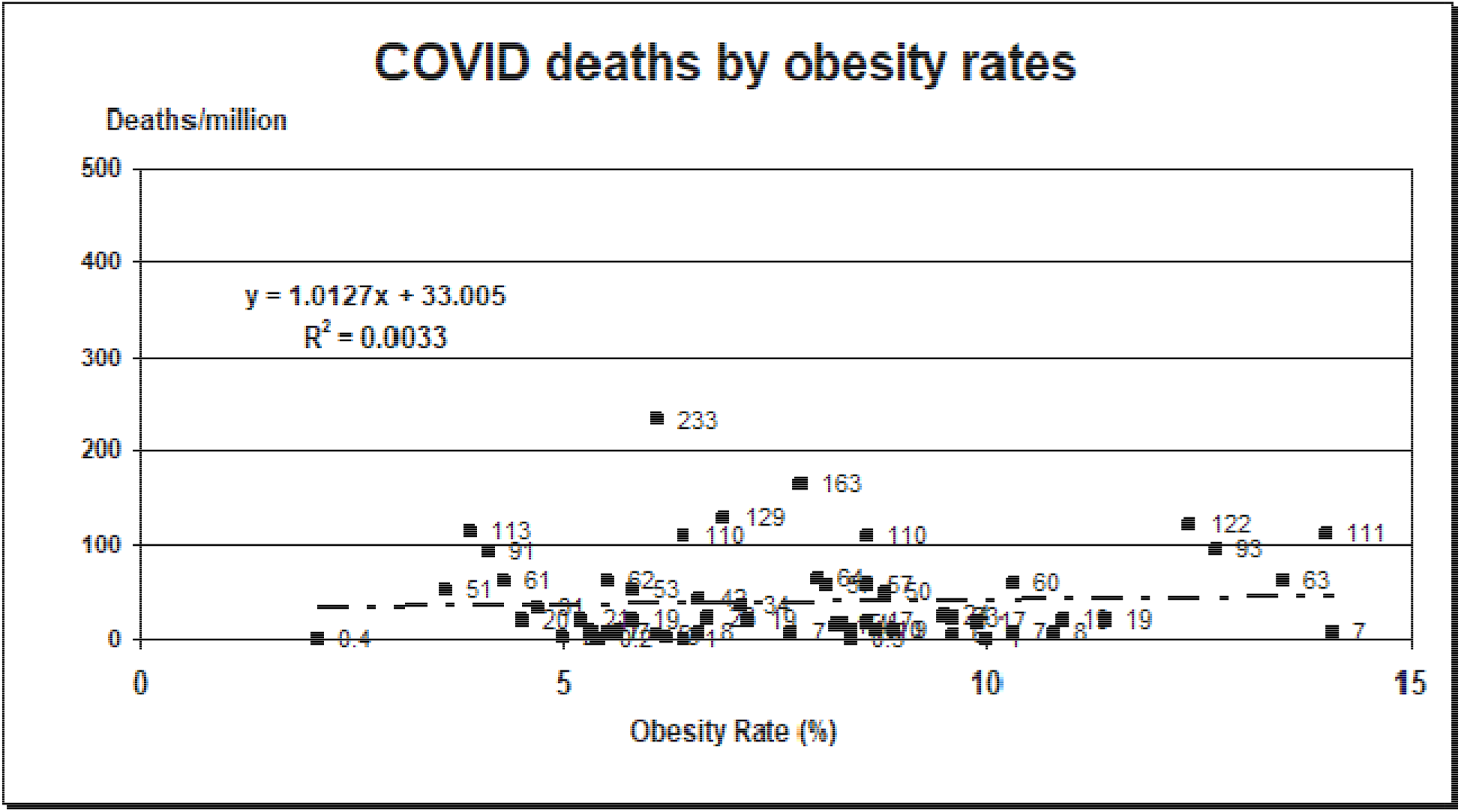
Association of 2016 obesity rates and COVID-19 deaths/million population for 57 countries with obesity rates <15%. Note different scale from Figure 1.

## 4. Discussion

### 4.1. Obesity matters

Results do not fully answer the question of how much of the difference in death rates is due to obesity, but they should make quite clear that obesity matters. With obesity rates < 15%, death rates are not associated with obesity and the slope of the fitted line=1. If all countries had obesity rates <15%, this study would be unnecessary. Starting with the 10 most populous countries and including all obesity rates, as more countries were added to the model based on population size, R^2^ values decreased from 0.9 to 0.3. That result may reflect the changing contribution from countries with obesity rates <15%. At the same time, the slope of the fitted line decreases by about half, from 51.0 to 27.9. Thus, for nearly 90% of the world population obesity appears to account for about one-third or more of the difference in COVID death rates. R^2^ drops to 0.2 when all 167 countries with data are included with little change in slope. Looking at Figure 1. suggests that attempting to estimate how many lives might have been saved if the obesity rate was reduced by a certain percentage will not be very precise. On the other hand, for countries with obesity rates above 15%, it is quite likely that their COVID death toll could have been reduced if their obesity rate were lower. And if the obesity rate could have been reduced to <15% it is relatively certain that COVID deaths would have been <200/million.

### 4.2. Relevance and questions

These results are consistent with data on hospitalizations and deaths from COVID-19 in the US (4) and worldwide (7). Not only is obesity one of the 6 underlying conditions (4), it is also a known risk factor for 4 others: asthma, cardiovascular disease, diabetes, and hypertension (8). Lowering the obesity rates in countries where it is >15% could potentially reduce deaths from COVID and from many chronic diseases (8). At least in the US, this would not be easy as obesity rates are currently increasing (9), but behavior change could potentially be successful (10). Other factors that might contribute to the COVID death rate such as racial, ethnic, and age differences, and health care capacity and quality, would likely be just as difficult to change and appear not to have much effect in countries with obesity rates <15%.

Study results also raise questions: Is there something special about obesity rates <15%? Why are there so few countries with obesity rates between 10% and 15%? Is some of the variability with higher obesity rates due to the presence of chronic conditions such as diabetes and hypertension related to obesity?

### 4.3. Limitations

There are several limitations to this study. Quality of the data for different countries may vary in how it was collected and what standards were used. Other factors may affect death rates including quality of the healthcare system, access to care, racial and ethnic disparities that lead to differences in developing COVID, presence of other underlying conditions in the population (including heart disease, hypertension, diabetes, and those not listed such as smoking), how the pandemic was handled in each country, legislative mandates, mask wearing practices, and geographic and cultural differences that might affect transmission. However, those factors do not appear to appreciably affect death rates among counties with obesity rates <15%.

### 4.4. Conclusions

For most of the world, at least one-third of the variation in COVID-19 death rates among countries appears to be due to obesity. By contrast, for 57 countries with obesity rates <15%, death rates are low and apparently not associated with obesity. More study is needed to understand those differences and other factors that might be involved. For countries with obesity rates >15%, COVID-19 deaths could have potentially been reduced with lower obesity rates. Although it is likely too late for COVID-19, prioritizing reducing obesity rates could lower deaths from future pandemics and reduce morbidity and mortality from chronic conditions for which obesity is a known risk factor (8,10).

## Data Availability

References 5 & 6 provide details for data

https://www.worldometers.info/coronavirus/#countries.

https://web.archive.org/web/20200630202020/https://www.cia.gov/library/publications/the-world-factbook/rankorder/2228rank.html

## Acknowledgements

No funding was received for this study.

The author declares no conflicts of interest.

## References

1. The Epidemiological Characteristics of an Outbreak of 2019 Novel Coronavirus Diseases (COVID-19) - China CCDC, February 17 2020.

2. Report of the WHO-China Joint Mission on Coronavirus Disease 2019 (COVID-19) [Pdf] - World Health Organization, Feb. 28, 2020

3. Garg S, Kim L, Whitaker M, O’Halloran A, Cummings C, Holstein R, et al. Hospitalization rates and characteristics of patients hospitalized with laboratory-confirmed coronavirus disease 2019—COVID-NET, 14 States, March 1–30, 2020. MMWR Morb Mortal Wkly Rep. 2020;69:458–64. PubMed https://doi.org/10.15585/mmwr.mm6915e3

4. Adams ML, Katz DL, Grandpre J. Updated estimates of chronic conditions affecting risk for complications from coronavirus disease, United States. Emerg Infect Dis. 2020 Sep [date cited]. https://doi.org/10.3201/eid2609.20211

5. Worldometer. COVID-19 Coronavirus Pandemic. Updated February 25, 2021. https://www.worldometers.info/coronavirus/#countries. Accessed February 25, 2021.

6. Central Intelligence Agency. The World Factbook. Country comparison: obesity - adult prevalence rate 2016. https://web.archive.org/web/20200630202020/https://www.cia.gov/library/publications/the-world-factbook/rankorder/2228rank.html. Accessed March 1, 2021.

7. Popkin BM, D. S, Green WD, et al. Individuals with obesity and COVID-19: A global perspective on the epidemiology and biological relationships. Obes Rev. 2020;21(11):e13128. doi:10.1111/obr.13128.

8. Adams M, Grandpre J, Katz D, Shenson D. The impact of key modifiable risk factors on leading chronic conditions. Prev Med. 2019 Mar;120:113–118. https://doi.org/10.1016/j.ypmed.2019.01.006.

9. Pernenkil V, Wyatt T, Akinyemiju T. Trends in smoking and obesity among US adults before, during, and after the great recession and Affordable Care Act roll-out. Prev Med. 2017 Sep;102:86–92. doi: 10.1016/j.ypmed.2017.07.001. Epub 2017 Jul 8.

10. Brownson RC, Remington PL, Wegner, MV. Chronic Disease Epidemiology and Control. 4th ed. Washington, DC: American Public Health Association; 2016.

